# UTILISATION AND DETERMINANTS OF BLOOD CULTURE IN MANAGING SEPSIS AMONG HOSPITALISED CHILDREN <5 YEARS: A MIXED-METHOD STUDY AT FOUR AMR SURVEILLANCE SITES IN UGANDA, 2024-2025

**DOI:** 10.64898/2026.03.03.26347550

**Authors:** Rogers Kisame, Ronald Kooko, Susan Nabadda, Ibrahim Mugerwa, Saudah Kizito Namubiru, Simon Kasango Dembe, Ceaser Nyolimati Adibaku, Anita Kisakye, Gideon Matovu, Henry Kajumbula, Joel Bazira, Wahab Kumakech Adubango, Philip Syagi Wandera, Enock Padere, Christopher Harold Amandu, Patricia Nahirya Ntege, Dithan Kiragga, Peter Elyanu

**Author notes:** Correspondence: Rogers Kisame.

## Abstract

Sepsis caused by drug-resistant pathogens remains a major contributor to under-five mortality in low- and middle-income countries, threatening progress toward Sustainable Development Goal (SDG) 3.2. Blood culture, the gold standard for sepsis diagnosis and antimicrobial stewardship, remains underutilised in routine pediatric care. This study assessed the extent and determinants of blood culture utilisation among hospitalised children under five years with suspected sepsis at four antimicrobial resistance (AMR) surveillance sites in Uganda. We conducted a cross-sectional mixed-methods study involving retrospective review of 384 pediatric patient records and in-depth interviews with 20 clinicians. Modified Poisson regression was used to identify factors associated with blood culture requests, while thematic analysis explored behavioral and contextual influences on diagnostic practices. Blood cultures were requested in 28.1% of suspected sepsis cases. Higher utilisation was independently associated with markers of clinical severity, including severe acute malnutrition (adjusted prevalence ratio [aPR] 1.3, 95% CI: 1.14–1.34), sickle cell disease (aPR 1.3, 95% CI: 1.19–1.40), and presence of WHO danger signs (aPR 1.1, 95% CI: 1.00–1.14). Senior clinician involvement (aPR 1.2, 95% CI: 1.08–1.32) and consultant review (aPR 1.4, 95% CI: 1.21–1.48) were also associated with higher use, while prior antibiotic exposure reduced the likelihood of blood culture request (aPR 0.9, 95% CI: 0.84–0.96). Qualitative findings identified four overarching themes influencing diagnostic behavior: motivation amid systemic constraints, institutional and environmental barriers, mentorship and teamwork, and emotional fatigue in the context of adaptive practices. Despite high clinician awareness, blood culture utilisation remains low, driven primarily by health system fragility, inefficient workflows, and emotional exhaustion rather than knowledge gaps. Improving utilisation will require integrated behavioral, workflow, and structural interventions, including clinical decision support and strengthened microbiology laboratory capacity, to enhance pediatric sepsis care, antimicrobial stewardship, and progress toward SDG 3.2.

## Introduction

Sepsis due to drug-resistant pathogens is a significant cause of childhood morbidity and mortality, particularly in low- and middle-income countries (LMICs) [1]. Globally, an estimated 2.9 million sepsis cases and 2.4 million sepsis-related deaths occur annually among children under five years of age [2]. In Uganda, children under five remain especially vulnerable to severe infections, and timely, accurate diagnosis is crucial to improving outcomes and curbing the growing threat of antimicrobial resistance (AMR). Reducing neonatal mortality from bacterial infections in LMICs is therefore critical to achieving Sustainable Development Goal (SDG) 3.2, which aims to reduce neonatal mortality to at least 12 per 1,000 live births by 2030 [3]. Without a significant reduction in neonatal mortality, meeting SDG goals remains unlikely.

Blood culture is the gold standard for diagnosing bloodstream infections and guiding targeted antibiotic therapy. Despite its diagnostic and antimicrobial stewardship value, blood culture utilisation in many resource-limited settings remains suboptimal. A prospective cohort study among neonates and young infants with suspected sepsis in Uganda found that only 11% of blood cultures yielded bacterial growth, and that Gram-negative isolates exhibited high resistance to first-line antibiotics [4]. Similarly, a 2021 cross-sectional study of blood culture in febrile children under five years at Uganda’s AMR surveillance sites reported a diagnostic yield of only 4.9%, highlighting significant challenges in laboratory practices and adherence to quality standards [5].

Routine AMR surveillance data from Uganda’s tertiary hospitals further highlight a considerable gap in blood culture utilisation. Between 2020 and 2023, isolates recovered from blood specimen accounted for only 8.9% of all bacterial pathogens reported by the national AMR surveillance network, suggesting persistent underutilization or systemic barriers to blood culture execution [6]. Previous reports attribute this to the patient costs, limited microbiology capacity, and recurrent supply-chain constraints [7]. Additionally, delayed feedback and limited clinician engagement undermine timely and accurate diagnosis, negatively impacting antimicrobial stewardship.

Beyond structural limitations, behavioral and contextual determinants influencing clinicians’ blood culture utilisation remain poorly understood. The Theoretical Domains Framework (TDF) offers a comprehensive behavioral lens for exploring these factors by integrating psychological, social, and environmental influences on healthcare decision-making [8]. The TDF explores domains such as knowledge, skills, and beliefs about consequences, professional role, environmental context, and reinforcement, making it well-suited to investigate the individual and contextual drivers of diagnostic decision-making in routine clinical care.

Despite substantial Government of Uganda investments in strengthening microbiology laboratory capacity, including blood culture through the National AMR surveillance programme, supported by partners like the UK Fleming Fund, including(i) the rollout of electronic laboratory information systems (A-LIS), (ii) expansion of regional microbiology testing services through workforce capacity building, provision of critical reagents, equipment procurement and services, and (iii) enhanced quality assurance, blood culture remains underutilized among pediatric inpatients. National AMR data show that only 8.9% of isolates originate from blood samples despite the high burden of sepsis in this age group [6]. This mismatch suggests that laboratory strengthening alone is insufficient to change clinical behavior unless behavioral and workflow factors are also addressed.

Understanding these behavioral determinants is particularly important given the Sustainable Development Goal (SDG) target 3.2. Optimising diagnostic utilisation in pediatric sepsis care aligns directly with this goal and with global AMR containment strategies.

Therefore, this study sought to examine the extent and determinants of blood culture utilisation in the management of sepsis among hospitalised children <5 years at four AMR surveillance sites in Uganda. Specifically, we assessed the rate of blood culture requests, identified clinical and contextual predictors of utilisation, and explored clinicians’ behavioral determinants using the Theoretical Domains Framework (TDF). Findings from this study will provide practical evidence to inform diagnostic stewardship interventions, enhance antimicrobial stewardship, ultimately improve patient outcomes and advance progress toward SDG 3.2.

## Materials and Methods

### Study design and setting

This study employed a cross-sectional mixed-methods retrospective design combining quantitative and qualitative approaches at four Regional Referral Hospitals (RRHs) enrolled in Uganda’s national antimicrobial resistance (AMR) surveillance network, purposively selected to ensure balanced geographic representation across Uganda’s Central, Western, Eastern, and Northern regions: Jinja RRH (Central region), Mbarara RRH (Western region), Mbale RRH (Eastern region), and Arua RRH (Northern region). Additionally, these sites served as demonstration units for implementing clinician engagement quality-improvement initiatives for enhanced antimicrobial stewardship, making them strategically relevant to the study. The quantitative component involved a retrospective review of pediatric inpatient records. In contrast, the qualitative component consisted of in-depth interviews guided by the Theoretical Domains Framework (TDF) with clinicians involved in the management of children with suspected sepsis. The mixed-methods approach enabled triangulation of utilisation patterns with clinicians’ behavioural and contextual experiences influencing blood culture use.

### Study population

The quantitative component involved reviewing clinical patient records of children under five years of age who were hospitalised with suspected sepsis at the four participating RRHs between September 2024 and September 2025. The qualitative component targeted clinicians directly involved in the clinical assessment, diagnosis, and management of pediatric patients with suspected sepsis on pediatric wards at the participating hospitals during the study period.

### Inclusion criteria

The quantitative component included clinical records of children aged under five years who were hospitalised with suspected sepsis at the four participating regional referral hospitals between September 2024 and September 2025. Records were eligible if the child had a documented clinical diagnosis of sepsis or suspected sepsis at admission and sufficient documentation on clinical presentation, admitting cadre, and investigations ordered during hospitalisation.

For the qualitative component, clinicians were eligible if they were directly involved in the clinical management of pediatric patients with suspected sepsis in the participating hospitals’ pediatric wards during the study period.

### Exclusion criteria

Patient records were excluded if the child was aged five years or older, if the diagnosis of sepsis was not documented, or if the clinical file lacked key information on investigations or treatment decisions.

Clinicians were excluded from the qualitative interviews if they were not directly involved in pediatric sepsis management, were on short-term rotation, or declined to provide informed consent.

### Sample size and sampling

Using the Kish-Leslie formula, with a prevalence of 50% (since the prevalence of blood culture utilisation among under-fives in the study setting was unknown) and a 5% margin of error, the sample size for the quantitative component was calculated at 384 patient files.

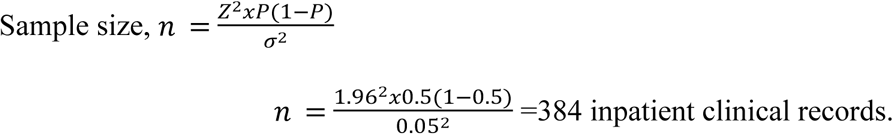

To ensure representativeness, sampling was done proportionate to the annual volume of blood culture tests performed at each site. Using probability proportionate to size (PPS), the following sample distribution was obtained:

**Table 1:** Sample showing sample size per site.

| Surveillance site | Sampling frame (average annual cultures) | Proportion of records for patients aged <5 years | Sample size |
| --- | --- | --- | --- |
| Mbarara RRH | 6,788 | 0.4438 | 170 |
| Jinja RRH | 3,204 | 0.2095 | 80 |
| Mbale RRH | 3,488 | 0.2280 | 88 |
| Arua RRH | 1,816 | 0.1187 | 46 |
| **Total** | **15,296** | **1** | **384** |

Using the patients’ files (children under five years old clinically diagnosed with sepsis) for September 2024 to September 2025 as the sampling frame and a systematic random sampling technique, participants were selected for the study until the estimated sample size was reached.

For the qualitative component, 20 clinicians (five per site) were purposively selected. Recruitment continued until data saturation [9], ensuring adequate representation of diverse clinical cadres and experiences across sites.

### Study variables

The primary outcome variable was blood culture utilisation, defined as whether a blood culture was requested during the child’s hospital admission for suspected sepsis (Yes/No).

Independent variables included the child’s age and gender, presence of fever (≥38°C), WHO danger signs, severe acute malnutrition, comorbidities, length of hospital stay, preadmission antibiotic use, day and time of admission, and admitting clinician cadre. The contextual factors were conceptualised based on the TDF and included clinician knowledge, skills, beliefs about consequences, professional role, environmental context, reinforcement, and social influences.

### Data collection

#### Patient record review

Data were collected over one month, from 1st September 2025 to 30th September 2025, using a structured data abstraction tool. The tool captured demographic information, clinical characteristics, admitting clinician cadre, timing of admission, preadmission antibiotic use, and whether a blood culture was requested during hospitalization.

#### Clinician interviews

Qualitative data were collected through face-to-face semi-structured interviews using an interview guide informed by the Theoretical Domains Framework (TDF). Interviews were conducted in private settings by trained qualitative researchers, audio-recorded with participant consent, and transcribed verbatim to ensure accuracy and completeness.

**Table 2:** TDF domain and interview guide questions.

| **TDF Domain** | **Key questions based on the TDF to assess behavioral and contextual influences on blood culture utilisation** |
| --- | --- |
| Knowledge | What do you know about the role, indications, and interpretation of blood cultures in managing sepsis among under-five children |
| Skills | What skills are needed to ensure a good-quality blood culture sample is collected in young children? |
| Social/professional role & identity | How does the use of blood cultures fit into your role? |
| Beliefs about capabilities | How confident are you in ensuring that blood cultures are consistently requested, collected, and results used in patient management? |
| Optimism | How optimistic are you that consistent use of blood cultures could improve sepsis outcomes among children? |
| Beliefs about consequences | What do you see as the benefits and drawbacks of using blood cultures in routine sepsis management? |
| Reinforcement | Have you or your team received any feedback or recognition regarding blood culture use or outcomes? |
| Intentions | When managing a child with suspected sepsis, do you usually intend to request a blood culture? |
| Goals | What are your main goals in managing sepsis in children, and how do blood cultures fit into those goals? |
| Memory, attention & decision processes | How do you remember or decide to order a blood culture during your rounds or consultations? |
| Environmental context & resources | What environmental or institutional factors affect blood culture use here? |
| Social Influences | How do colleagues or caregivers influence your decision to use blood cultures? |
| Emotion | What feelings do you experience when requesting or performing blood cultures in young children? |
| Behavioural Regulation | What systems or reminders exist to ensure blood cultures are ordered, and results are acted upon? |

#### Data analysis

Data analysis was performed using Statistical Package for Social Sciences (SPSS) version 26. Descriptive statistics were used to summarise participants’ characteristics, with categorical variables presented as frequencies and percentages. At the bivariate level, a Modified Poisson regression with robust standard errors was used to identify factors associated with blood culture requests, yielding crude prevalence ratios and P-values at 95% confidence intervals. Variables with a p-value < 0.20 at the bivariate level were retained for inclusion in the multivariable Modified Poisson regression to avoid excluding potential confounders. Multivariable Modified Poisson regression analysis was then performed to identify factors independently associated with blood culture request, and adjusted prevalence ratios (aPRs) with corresponding 95% confidence intervals were reported. Statistical significance was set at p < 0.05.

Audio-recorded interviews were transcribed verbatim and analysed using thematic analysis informed by the Theoretical Domains Framework (TDF). The research team familiarised themselves with the data, applied open coding in NVivo version 15, and mapped emerging codes deductively onto eight TDF domains to capture behavioural determinants of blood culture utilisation. Through iterative comparison and team discussion, related codes were grouped into analytical categories that described patterns across cadres and facilities. These categories were then synthesised into four overarching themes that reflected clinicians’ experiences and behavioural drivers: motivation amidst systemic limitations; environmental and institutional barriers; mentorship, teamwork, and social reinforcement; and emotional fatigue and adaptive behaviour. Rigour was ensured through coder consensus, triangulation across cadres, and maintenance of an audit trail of analytics.

#### Ethical considerations

This study was reviewed and approved by the Makerere University School of Public Health Research and Ethics Committee under reference number MakSPH-REC_506. Additionally, administrative clearance was obtained from the management of each participating Regional Referral Hospital (RRH).

For the quantitative component, only secondary data were used; no direct patient contact occurred. Patient identifiers were removed, and unique study codes were used to ensure confidentiality. For the qualitative interviews, verbal informed consent was obtained from all clinician participants before data collection. Participation was voluntary, and respondents could withdraw at any point without consequence. All electronic data were password-protected and stored on secure servers accessible only to the research team. The study posed minimal risk but offers substantial public health benefits through evidence to enhance pediatric sepsis management and antimicrobial stewardship in Uganda.

## Results

### Sociodemographic characteristics of the respondents

A total of 384 pediatric patient records from four AMR surveillance sites were reviewed. The mean age was 14.7 ± 16.9 months, with a median age of 7 months (IQR 3–23). More than half of the participants (58.9%) were aged below 12 months, and 52.3% were female. Nearly half (44.3%) were admitted at Mbarara Regional Referral Hospital, followed by Mbale (22.9%), Jinja (20.8%), and Arua (12.0%).

Most children (87.5%) were not referred from another health facility. Fever at admission (≥38°C) was documented in 57.8% of cases, while 65.1% presented with at least one WHO danger sign. Regarding length of hospital stay, 46.9% of children were admitted for 4–7 days, 26.0% for 0–3 days, and 27.1% for eight days or more. Severe acute malnutrition was documented in 4.9% of cases, and 8.9% had sickle cell disease. Most admissions occurred during the daytime (78.4%) and on weekdays (82.3%). (Table 3)

**Table 3:** Socio-demographic characteristics of under-five children with sepsis at four AMR surveillance sites (N = 384)

| Variables | Frequency (n) | Percentage (%) |
| --- | --- | --- |
| **Age (Months)** |  |  |
| Age (Mean ± SD) | 14.7±16.9 | |
| Age (Medium, IQR) | 7, (3, 23) |  |
| <12 | 226 | 58.9 |
| 12-35 | 92 | 24.0 |
| $\geq 36$ | 66 | 17.2 |
| **Gender** |  |  |
| Female | 201 | 52.3 |
| Male | 183 | 47.7 |
| **Surveillance site** |  |  |
| Arua RRH | 46 | 12.0 |
| Jinja RRH | 80 | 20.8 |
| Mbale RRH | 88 | 22.9 |
| Mbarara RRH | 170 | 44.3 |
| **Referral status** |  |  |
| No | 336 | 87.5 |
| Yes | 48 | 12.5 |
| **Temp at admission $\geq 38$** |  |  |
| No | 162 | 44.2 |
| Yes | 222 | 57.8 |
| **Presence of WHO danger signs at admission.** |  |  |
| No | 134 | 34.9 |
| Yes | 250 | 65.1 |
| **Length of Hospital stay (days)** |  |  |
| 0-3 | 100 | 26.0 |
| 4-7 | 180 | 46.9 |
| ≥38 | 104 | 27.1 |
| **Sickle cell status** |  |  |
| No | 350 | 91.1 |
| Yes | 34 | 8.9 |
| **Severe acute malnutrition** |  |  |
| No | 365 | 95.1 |
| Yes | 19 | 4.9 |
| **Time of admission** |  |  |
| Night | 83 | 21.6 |
| Day | 301 | 78.4 |
| **Date of admission** |  |  |
| Week end | 68 | 17.7 |
| Week day | 316 | 82.3 |

### Level of utilisation of Blood culture in the management of sepsis

Of the 384 patients, 108 (28.1%) had blood culture requests (Figure 1).

**Figure 1:**
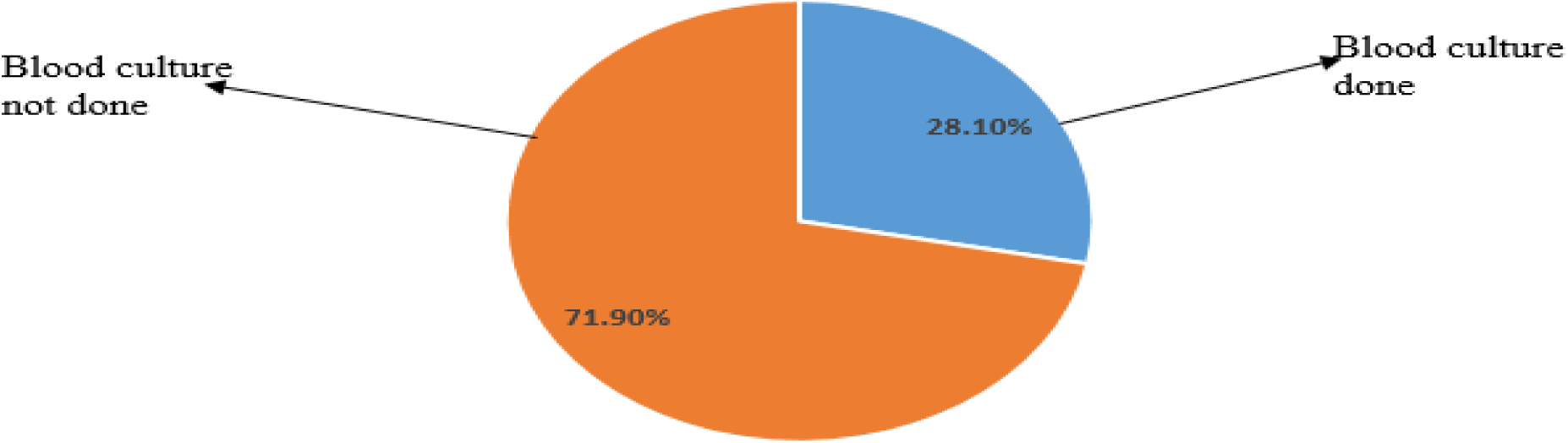
Level of Blood culture utilisation.

### Factors associated with Blood culture utilisation in the management of sepsis in children under five at AMR surveillance sites

At multivariable analysis, several factors were independently associated with blood culture utilisation.

Children aged 12–35 months (aPR = 0.9, 95% CI 0.84–0.94, p < 0.001) and those aged ≥36 months (aPR = 0.9, 95% CI 0.81–0.92, p < 0.001) were less likely to have a blood culture requested compared with children aged <12 months. Blood culture utilisation was higher among children admitted by more senior clinicians. Admission by a senior house officer or medical officer (aPR = 1.2, 95% CI 1.08–1.32, p = 0.001) and by a consultant (aPR = 1.4, 95% CI 1.21–1.48, p < 0.001) was associated with a higher likelihood of blood culture request compared with admission by a nurse. Clinical severity and comorbidity indicators were also associated with blood culture utilisation. Children with severe acute malnutrition (aPR = 1.3, 95% CI 1.14–1.34, p < 0.001), sickle cell disease (aPR = 1.3, 95% CI 1.19–1.40, p < 0.001), fever at admission (≥38°C) (aPR = 1.2, 95% CI 1.10–1.25, p < 0.001), and WHO danger signs (aPR = 1.1, 95% CI 1.00–1.14, p = 0.046) were more likely to have a blood culture requested. Longer hospital stay (≥8 days) (aPR = 1.1, 95% CI 1.01–1.16, p = 0.012) was associated with increased culture utilisation. Timing of admission was significantly associated with blood culture use. Children admitted during the daytime (aPR = 1.1, 95% CI 1.04–1.19, p = 0.001 and on weekdays (aPR = 1.1, 95% CI 1.02–1.17, p = 0.008) were more likely to have a blood culture requested compared with those admitted at night or during weekends. Contrary to preadmission, antibiotic use was associated with a lower likelihood of blood culture request (aPR = 0.9, 95% CI 0.84–0.96, p = 0.001). (Table 4)

**Table 4.**
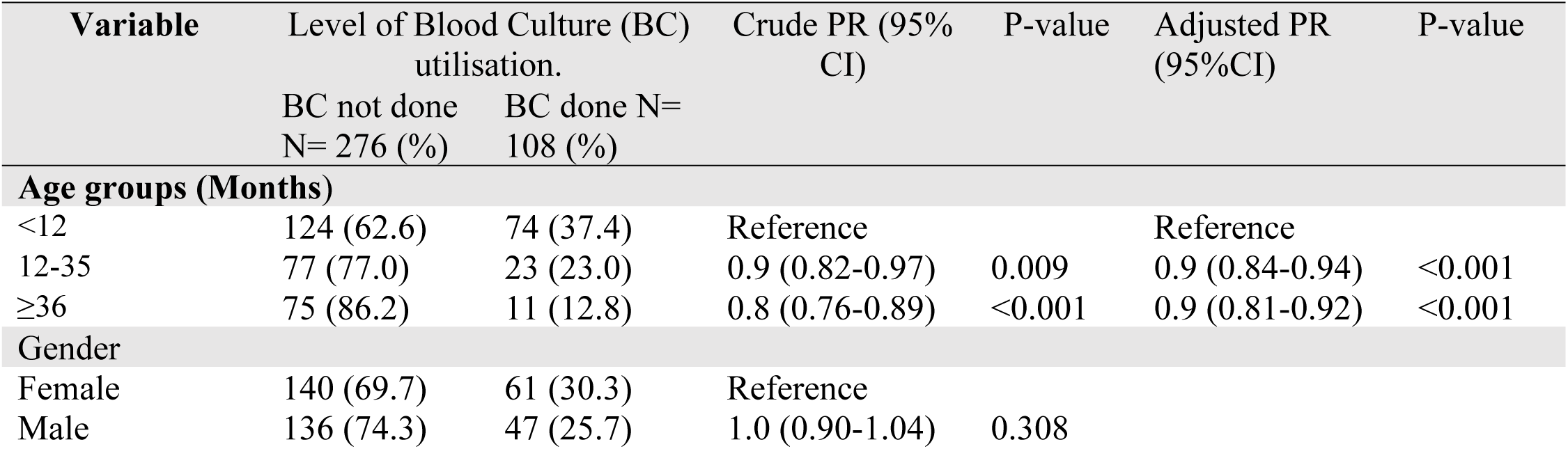
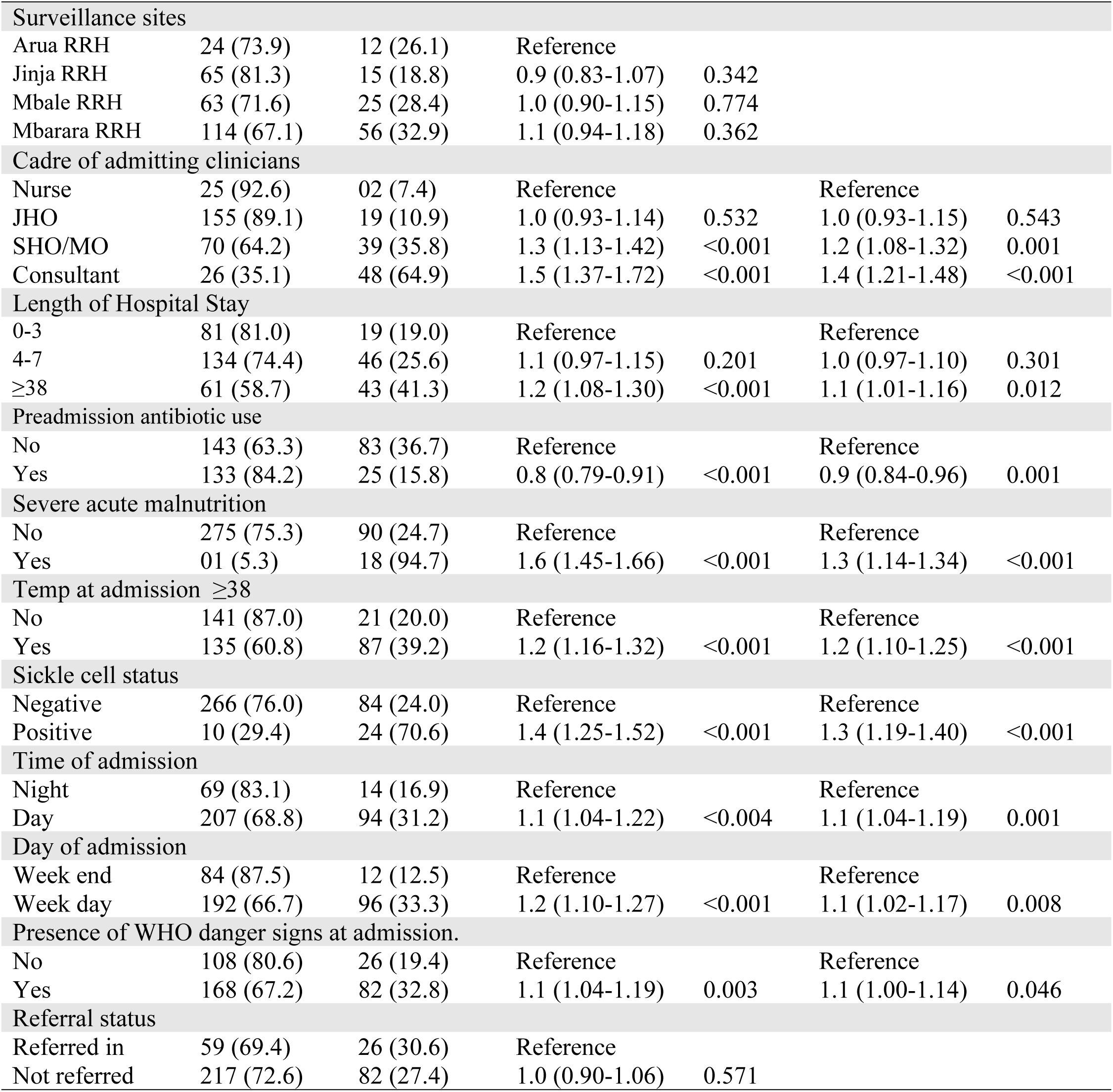
Factors associated with Blood culture utilisation in the management of sepsis in children under five.

| Variable | Level of Blood Culture (BC) utilisation. |  | Crude PR (95% CI) | P-value | Adjusted PR (95%CI) | P-value |
| --- | --- | --- | --- | --- | --- | --- |
|  | BC not done<br>N= 276 (%) | BC done N=<br>108 (%) |  |  |  |  |
| Age groups (Months) |  |  |  |  |  |  |
| <12 | 124 (62.6) | 74 (37.4) | Reference |  | Reference |  |
| 12-35 | 77 (77.0) | 23 (23.0) | 0.9 (0.82-0.97) | 0.009 | 0.9 (0.84-0.94) | <0.001 |
| ≥36 | 75 (86.2) | 11 (12.8) | 0.8 (0.76-0.89) | <0.001 | 0.9 (0.81-0.92) | <0.001 |
| Gender |  |  |  |  |  |  |
| Female | 140 (69.7) | 61 (30.3) | Reference |  |  |  |
| Male | 136 (74.3) | 47 (25.7) | 1.0 (0.90-1.04) | 0.308 |  |  |

|  |  |  |  |  |  |  |
| --- | --- | --- | --- | --- | --- | --- |
| **Surveillance sites** |  |  |  |  |  |  |
| Arua RRH | 24 (73.9) | 12 (26.1) | Reference |  |  |  |
| Jinja RRH | 65 (81.3) | 15 (18.8) | 0.9 (0.83-1.07) | 0.342 |  |  |
| Mbale RRH | 63 (71.6) | 25 (28.4) | 1.0 (0.90-1.15) | 0.774 |  |  |
| Mbarara RRH | 114 (67.1) | 56 (32.9) | 1.1 (0.94-1.18) | 0.362 |  |  |
| **Cadre of admitting clinicians** |  |  |  |  |  |  |
| Nurse | 25 (92.6) | 02 (7.4) | Reference |  | Reference |  |
| JHO | 155 (89.1) | 19 (10.9) | 1.0 (0.93-1.14) | 0.532 | 1.0 (0.93-1.15) | 0.543 |
| SHO/MO | 70 (64.2) | 39 (35.8) | 1.3 (1.13-1.42) | <0.001 | 1.2 (1.08-1.32) | 0.001 |
| Consultant | 26 (35.1) | 48 (64.9) | 1.5 (1.37-1.72) | <0.001 | 1.4 (1.21-1.48) | <0.001 |
| **Length of Hospital Stay** |  |  |  |  |  |  |
| 0-3 | 81 (81.0) | 19 (19.0) | Reference |  | Reference |  |
| 4-7 | 134 (74.4) | 46 (25.6) | 1.1 (0.97-1.15) | 0.201 | 1.0 (0.97-1.10) | 0.301 |
| ≥38 | 61 (58.7) | 43 (41.3) | 1.2 (1.08-1.30) | <0.001 | 1.1 (1.01-1.16) | 0.012 |
| **Preadmission antibiotic use** |  |  |  |  |  |  |
| No | 143 (63.3) | 83 (36.7) | Reference |  | Reference |  |
| Yes | 133 (84.2) | 25 (15.8) | 0.8 (0.79-0.91) | <0.001 | 0.9 (0.84-0.96) | 0.001 |
| **Severe acute malnutrition** |  |  |  |  |  |  |
| No | 275 (75.3) | 90 (24.7) | Reference |  | Reference |  |
| Yes | 01 (5.3) | 18 (94.7) | 1.6 (1.45-1.66) | <0.001 | 1.3 (1.14-1.34) | <0.001 |
| **Temp at admission ≥38** |  |  |  |  |  |  |
| No | 141 (87.0) | 21 (20.0) | Reference |  | Reference |  |
| Yes | 135 (60.8) | 87 (39.2) | 1.2 (1.16-1.32) | <0.001 | 1.2 (1.10-1.25) | <0.001 |
| **Sickle cell status** |  |  |  |  |  |  |
| Negative | 266 (76.0) | 84 (24.0) | Reference |  | Reference |  |
| Positive | 10 (29.4) | 24 (70.6) | 1.4 (1.25-1.52) | <0.001 | 1.3 (1.19-1.40) | <0.001 |
| **Time of admission** |  |  |  |  |  |  |
| Night | 69 (83.1) | 14 (16.9) | Reference |  | Reference |  |
| Day | 207 (68.8) | 94 (31.2) | 1.1 (1.04-1.22) | <0.004 | 1.1 (1.04-1.19) | 0.001 |
| **Day of admission** |  |  |  |  |  |  |
| Week end | 84 (87.5) | 12 (12.5) | Reference |  | Reference |  |
| Week day | 192 (66.7) | 96 (33.3) | 1.2 (1.10-1.27) | <0.001 | 1.1 (1.02-1.17) | 0.008 |
| **Presence of WHO danger signs at admission.** |  |  |  |  |  |  |
| No | 108 (80.6) | 26 (19.4) | Reference |  | Reference |  |
| Yes | 168 (67.2) | 82 (32.8) | 1.1 (1.04-1.19) | 0.003 | 1.1 (1.00-1.14) | 0.046 |
| **Referral status** |  |  |  |  |  |  |
| Referred in | 59 (69.4) | 26 (30.6) | Reference |  |  |  |
| Not referred | 217 (72.6) | 82 (27.4) | 1.0 (0.90-1.06) | 0.571 |  |  |

### Qualitative Findings: Clinicians’ Perspectives on Blood Culture Utilisation

Twenty clinicians involved in pediatric sepsis management participated in the qualitative interviews. Participants included consultants, pediatric residents, medical officers, clinical officers, and nurses across the four AMR surveillance sites. Thematic analysis, guided by the Theoretical Domains Framework (TDF), identified four overarching themes capturing behavioural, institutional, and emotional factors influencing blood culture utilisation: (1) motivation amidst systemic limitations, (2) environmental and institutional barriers, (3) mentorship, teamwork, and social reinforcement, and (4) emotional fatigue and adaptive behaviour.

### THEME 1. Motivation amidst systemic limitations

Across all cadres, clinicians recognised blood culture as a critical diagnostic tool for confirming sepsis and guiding antimicrobial therapy, particularly in neonates and severely ill children. Participants frequently described blood cultures as the “gold standard” for diagnosing sepsis. *“Blood culture is, without doubt, the most important test when managing sepsis. It gives us a definitive diagnosis, tells us the exact bacteria and which antibiotic works.” (Consultant Paediatrician, KII_10).* Despite this strong motivation, clinicians reported that delayed turnaround times and limited feedback reduced the perceived clinical utility of blood culture results. “*Results come back after two or three days when the child is already discharged or improving. We end up treating blindly.” (Medical Officer, KII_4)*

Several participants described a sense of helplessness, knowing what should be done, but constrained by system inefficiencies. *“Even when you know it’s important, you lose morale when results are delayed or never come back.” (Clinical Officer, KII_19)*

### THEME 2. Environmental and institutional barriers

Environmental and institutional constraints were consistently described as significant barriers to blood culture utilisation. Participants reported recurrent stock-outs of culture bottles and reagents, laboratory understaffing, intermittent power supply, and delayed sample transport. *“Sometimes the lab tells us there are no bottles. We have to start antibiotics without doing the test.” (Paediatric Ward in-Charge Nurse, KII_5).* The consultant noted that these failures eroded the influence of clinical leadership. *“You can’t insist that juniors follow procedure when the system itself isn’t functioning. It’s demoralising.” (Consultant Paediatrician, KII_15)*

Cost-related barriers were also reported, particularly during periods of stock-out, when families were asked to seek blood culture services outside the hospital. *“When parents are told to buy bottles or have the culture done outside and they can’t, we just manage empirically. It’s heartbreaking.” (Senior house officer (SHO), KII_1).* These constraints undermined the integration of diagnostic stewardship into routine sepsis care.

### THEME 3. Mentorship, Teamwork, and Social Reinforcement

Clinicians highlighted the roles of leadership, mentorship, and collaboration between clinical and laboratory teams in promoting the use of blood cultures. Participants reported that mentorship initiatives and regular feedback improved accountability and encouraged consistent practice. *“During the antimicrobial resistance (AMR) mentorship, we used to get monthly reports. Those reports were eye-opening — they showed us the common organisms, resistance patterns, and contamination rates. It helped us see where we were improving or falling behind” (Consultant Paediatrician, KII_10).* Team cohesion also mattered. *“When consultants and lab staff work together, things move faster. Results come back on time, and people see the benefit.” ( (SHO), KII_6)*

However, participants noted that when mentorship activities or feedback mechanisms were discontinued, adherence to blood culture practices declined. *“Once mentorship stopped, the culture of collecting samples also faded.” (Clinical Officer, KII_9)*.

### THEME 4. Emotional fatigue and adaptive behaviour

Clinicians described a complex mix of stress, frustration, and resilience in navigating daily diagnostic challenges. *“It is nerve-wracking drawing blood from small babies, especially when parents are anxious. But it is part of what we do.” (Paediatric Nurse, KII_8)*

Repeated system failures bred disillusionment. *“After several times of doing everything right and still not getting results, you start to lose hope.” (Medical Officer, KII_3)*

Nevertheless, clinicians reported renewed motivation when blood culture results contributed to improved clinical outcomes, reinforcing the perceived value of diagnostic testing. *“When a culture result changes management and the child recovers, you remember why this test matters.” (Medical officer, KII_13)*.

### Integration of quantitative and qualitative findings

The quantitative findings demonstrated low overall blood culture utilisation, with higher use among children admitted by senior clinicians, those presenting during the day or on weekdays, and those with markers of clinical severity. The qualitative findings provided contextual insights into these patterns, describing how leadership, staffing availability, laboratory functionality, and emotional burden shaped clinicians’ diagnostic practices. Both findings illustrate how blood culture utilisation in pediatric sepsis care is influenced by the interplay among clinical severity, health system functionality, team dynamics, and individual clinician experience.

## Discussion

This mixed-methods study examined the extent and determinants of blood culture utilisation in the management of sepsis among hospitalised children under five years across four AMR surveillance sites in Uganda. Despite the established diagnostic and stewardship value of blood cultures, less than one-third of pediatric sepsis admissions had a culture requested. The findings highlight a striking gap between clinicians’ knowledge and the actual practice of diagnostic stewardship, reflecting both behavioral and systemic barriers that undermine rational antimicrobial use in resource-limited settings.

### Low utilisation of blood cultures despite clinician awareness

The overall utilisation rate of 28% observed in this study mirrors patterns reported elsewhere in sub-Saharan Africa, where resource shortages and workflow constraints limit the adoption of microbiological diagnostics in routine care [8, 9]. Although clinicians recognised blood cultures as the “gold standard” for guiding antibiotic therapy, their use was frequently deprioritised due to delayed turnaround times and logistical failures. This difference between clinical intent and implementation has also been documented in Kenya and Nigeria, where delayed feedback and limited laboratory capacity discouraged sample collection [10, 11]. The consistency of this finding across settings suggests that strengthening laboratory clinical linkages is as critical as expanding infrastructure.

### Influence of clinician cadre and leadership

The study revealed that blood culture requests were notably higher among children managed by consultants and senior medical officers than those attended to by junior clinicians or nurses.

Qualitative findings further illustrated that senior clinicians often model evidence-based practice, mentor junior staff, and set expectations that reinforce diagnostic stewardship. This leadership effect shapes clinical norms and fosters accountability in test ordering and result follow-up, creating a culture of diagnostic vigilance. In contrast, wards with limited senior oversight tended to exhibit inconsistent blood culture utilisation, reflecting how hierarchical structures and role modelling influence adherence to diagnostic best practices.

Evidence from other settings supports these observations. A multicenter study in the United States found that leadership engagement and structured feedback loops significantly improved clinicians’ compliance with diagnostic protocols [12]. Similar findings from a study in Malawi highlight that sustained mentorship, supervision, and recognition by senior clinicians enhance stewardship behaviors and adherence to sepsis management guidelines [13]. Strengthening clinical leadership and institutionalising feedback mechanisms could therefore serve as key levers for improving the quality and consistency of microbiological practices in pediatric care.

### Impact of systemic and institutional barriers

Systemic and institutional constraints were identified as significant blood determinants of cultural utilisation. Clinicians frequently cited stock-outs of bottles and reagents, intermittent power supply, limited laboratory staffing, and delayed specimen transport as recurring challenges. These barriers were particularly evident during weekends and night shifts, when laboratory support was minimal. Similar operational gaps have been documented in Uganda’s national AMR surveillance reports and the WHO GLASS findings, which highlight infrastructure and supply-chain fragility as key obstacles to reliable microbiological diagnostics [4, 14]. Such deficiencies not only restrict access to essential testing but also create frustration and diagnostic fatigue among clinicians, reinforcing reliance on empirical therapy rather than evidence-based care. Overcoming these systemic barriers requires coordinated investment in health system strengthening. Ensuring uninterrupted supply of culture consumables, maintaining functional power backup systems, and establishing 24-hour laboratory coverage would significantly improve diagnostic reliability. In addition, creating clear communication and escalation protocols for urgent pediatric sepsis samples can enhance turnaround time and clinician trust in laboratory results. These interventions align with global recommendations for diagnostic stewardship and AMR containment, emphasising that functional laboratory infrastructure is foundational to rational antimicrobial use in low-resource hospital settings [14, 5].

### Clinical severity, comorbidity, and diagnostic prioritisation

Children presenting with severe acute malnutrition, sickle cell disease, or WHO danger signs were more likely to have blood cultures requested. This pattern indicates that clinicians tend to prioritise microbiological testing for children perceived to be at the highest risk of severe infection. Such diagnostic prioritisation reflects rational triage in high-pressure environments where both staff and laboratory resources are constrained. Similar findings have been reported in Uganda and Ghana, where clinicians relied on severity cues, such as shock, altered consciousness, or failure to improve with empirical therapy, to decide when to request cultures [15, 16]. This approach, though pragmatic, may inadvertently exclude children with early or atypical presentations of sepsis, leading to delayed or missed diagnoses.

Emerging evidence underscores that consistent use of diagnostic tools, rather than selective ordering, improves sepsis outcomes and supports antimicrobial stewardship [8]. Introducing structured prompts for blood culture requests within pediatric admission or sepsis management forms could help standardise practice and reduce missed opportunities for diagnosis. Reinforcing these prompts through mentorship, checklists, or electronic systems would support clinical decision-making without adding substantial workload. Such integration could balance the need for clinical judgment with the goal of equitable access to diagnostics for all pediatric patients with suspected sepsis.

### Preadmission antibiotic use and perceived diagnostic futility

The study found an inverse association between preadmission antibiotic use and blood culture requests, reflecting clinicians’ belief that prior antimicrobial exposure reduces the likelihood of positive culture results. This perception, while partly evidence-based, often leads to deferred or omitted culture testing. Similar views have been reported in Uganda and Ghana, where clinicians described blood culture as “futile” once empirical treatment had begun, particularly when laboratory feedback was delayed or inconsistent [17, 5]. Such perceptions are reinforced by repeated negative or contaminated test results, which erode confidence in the practical utility of microbiological testing in routine care.

Emerging evidence, however, shows that meaningful culture results can still be obtained in pre-treated patients, especially when samples are collected before multiple antibiotic doses or processed under optimised laboratory conditions [16, 18]. Addressing misconceptions about diagnostic futility requires ongoing education and effective communication between clinicians and laboratory personnel regarding optimal timing, sample handling, and result interpretation. Structured feedback mechanisms, such as regular laboratory-clinician meetings or shared dashboards, could help restore trust in culture results and promote more consistent utilisation, even among patients who have already received antibiotics.

### Emotional fatigue and adaptive behavior

The qualitative findings illuminated the emotional burden of diagnostic practice in resource-limited settings. Clinicians described frustration, helplessness, and fatigue arising from recurrent system failures such as stock-outs, delayed results, and unacknowledged efforts. Yet, they also expressed professional satisfaction when culture results guided recovery, demonstrating a spectrum of emotional responses that shaped adaptive behaviors ranging from disengagement to renewed motivation depending on institutional support. This pattern aligns with broader evidence linking emotional exhaustion to diminished diagnostic engagement in low-resource hospitals [19, 13]. A multicountry study in Ghana, Tanzania, Uganda, and Zambia, as well as similar studies from Tanzania and Kenya, have reported that high workloads, limited feedback, and inadequate supervision contribute to burnout and reduced adherence to diagnostic and stewardship protocols [20]. Conversely, supportive environments foster resilience and sustained diagnostic commitment. Studies from Malawi, Ghana, and Uganda show that mentorship, feedback, and professional recognition strengthen clinicians’ morale and improve adherence to sepsis and antimicrobial stewardship guidelines [8]. These findings suggest that addressing emotional fatigue requires systemic action beyond individual coping strategies. Integrating psychosocial support, regular feedback, and recognition for good diagnostic practice into hospital quality improvement programs could enhance clinician wellbeing and diagnostic performance [13, 14]. Promoting emotional resilience is therefore an essential component of sustaining diagnostic stewardship in high-pressure pediatric settings.

### Implications for policy and practice

Together, these findings demonstrate that low blood culture utilisation in Ugandan pediatric wards is not primarily a knowledge issue but a systemic functionality and behavioural reinforcement issue. Interventions must therefore operate on multiple levels: ensuring continuous supply of consumables and reliable laboratory operation, embedding reminders or checklists into sepsis care workflows, providing continuous mentorship and feedback, and creating collaborative ward–laboratory relationships. These interventions are consistent with WHO recommendations on diagnostic stewardship and AMR containment [14]. Strengthening these systems would not only enhance diagnostic accuracy but also directly contribute to Uganda’s AMR surveillance goals and to Sustainable Development Goal 3.2, which aims to end preventable child deaths.

### Strengths and limitations

This study’s strengths include its multicenter design across four AMR surveillance hospitals, the integration of quantitative and qualitative data, and the use of real-world hospital records. However, limitations include reliance on retrospective record reviews, which may have incomplete data, and the cross-sectional design, which precludes causal inference. The clinician interviews were limited to regional referral hospitals and may not reflect practices at lower-level or private facilities.

## Conclusion

Blood culture utilisation among under-five children with sepsis in Ugandan AMR surveillance hospitals remains low despite high clinician awareness. Practice is constrained by systemic barriers, workflow disruptions, and emotional fatigue, rather than by knowledge gaps. Sustainable improvement will require context-specific diagnostic stewardship interventions that combine infrastructural investment, leadership engagement, and behavioral reinforcement to strengthen rational antimicrobial use in pediatric care.

## List of abbreviations and acronyms

AMR: Antimicrobial Resistance
TDF: Theory Domain Framework
SSA: Sub-Saharan Africa
PPS: Probability Proportionate to Size
RRH: Regional Referral Hospital
WHO: World Health Organisation
PR: Prevalence Ratio
APR: Adjusted Prevalence Ratio
SHO: Senior House Officer

## Declarations

### Ethics approval and consent to participate

Ethical approval was sought from the Research and Ethics Committee of Makerere University School of Public Health. The study was conducted in accordance with the ethical principles of the Declaration of Helsinki. Permission to conduct the study was sought from the participants. The study’s purpose was explained, and the information obtained was used solely for research. Verbal informed consent was obtained from study participants before conducting any interviews, and confidentiality was ensured throughout the study. Administrative clearance was also sought from surveillance sites. All information provided by the respondents was confidential.

## Availability of data and materials

The data sets supporting the findings are readily available and have been submitted to the journal.

## Competing interests

The authors have declared that no competing interests exist.

## Consent for publication

Not applicable.

## Funding

The authors received no specific funding for this work.

## Author contributions

R K and R K conceived the study, designed the research, coordinated the surveillance activities, and drafted the initial manuscript. R K collected, cleaned, and analysed the data, developed the visualisations, and wrote the results section of the manuscript. P E and R K provided technical oversight, contributed to manuscript revisions, and ensured alignment with national AMR surveillance standards. R K, S K D, A K and G M supported data extraction and management and provided technical input on data analysis procedures. S N, I M, S K N, J B and H K supervised the laboratory components, reviewed microbiological methods, and provided expert guidance during study design and data interpretation. S K D, C N A and A K contributed to data collection, verification, and interpretation of site-level findings. R K, D K and P E provided overall project oversight, critically reviewed the manuscript, and guided the framing of public health implications. W K A, P S W, E P, and C H A contributed to site coordination, data acquisition, contextual interpretation of findings, and critical review of the manuscript. P N N reviewed the manuscript to ensure clarity, rigour, coherence, and completeness.

All authors reviewed, provided critical revisions, and approved the final manuscript for submission.

## Data Availability

The data sets supporting the findings are readily available and have been submitted to the journal under supporting information.

## Acknowledgements

We acknowledge the four surveillance sites that participated in this study: Mbarara RRH, Mbale RRH, Jinja RRH and Arua RRH.

## Supporting Information

**A S1 File. Quantitative dataset used for analysis.**

An anonymised dataset containing variables extracted from pediatric inpatient records used in the quantitative analysis of blood culture utilisation among children under five years with suspected sepsis at four AMR surveillance sites in Uganda.

**A S2 File. Qualitative interview transcripts.**

De-identified verbatim transcripts of key informant interviews conducted with clinicians involved in the management of pediatric sepsis were used for the qualitative component of the study.

## Notes

### Competing Interest Statement

The authors have declared no competing interest.

### Author Declarations

Research and Ethics Committee of Makerere University School of Public Health.

